# Argentine Registry of neurological manifestations due to coronavirus-19 (COVID-19)

**DOI:** 10.1101/2021.03.19.21253558

**Authors:** Lucas Alessandro, Franco Appiani, Mariana Bendersky, Brenda Borrego Guerrero, Guadalupe Bruera, Patricia Cairola, Ismael Calandri, Juan Martín Cardozo Oliver, María Emilia Clément, Marianna Di Egidio, José Luis Di Pace, Melina Diaconchuk, Celeste Esliman, Ma Martha Esnaola y Rojas, Julián Fernández Boccazzi, Andrea Fabiana Franco, Gisella Gargiulo, Daniela Laura Giardino, César Gómez, Ana Karina Guevara, Natalia Gutierrez, Javier Hryb, Ibarra Viviana, Franco Janota, Mabel Laserna, Luis Alfredo Larcher, Fernando Leone, Geraldine Luetic, Claudia Andrea Medina, María Laura Menichini, Gonzalo Nieto, María Fernanda Páez, Francisco Peñalver, Mónica Perassolo, Gabriel Persi, Claudia Pestchanker, Oscar Porta, Roberto Daniel Rey, Gabriel Eduardo Rodríguez, Marina Romano, Marcelo Rugiero, Patricia Saidón, María Florencia Sica, Erica Stankievich, Adriana Tarulla, Guillermo Zalazar

**Affiliations:** FLENI-; Hospital Universitario Fundación Favaloro; Universidad de Buenos Aires; IADIN; Hospital Italiano de Buenos Aires; Sanatorio Tandil; Hospital Privado de Rosario; Laboratorio Max Planck; Hospital CG Durand; Sanatorio Finochietto; Hospital Privado de la Comunidad; Hospital E. Tornú; Hospital San Luis; Hospital César Milstein; Sanatorio de la Trinidad Mitre; Hospital J.M. Ramos Mejía; Hospital Universitario CEMIC; CONICET; Mutual Rural ART; Hospital Enrique Vera Barros; Sanatorio Julio Méndez; Hospital Francisco Muñiz; Sanatorio del Norte; Centro Médico Roca; Instituto de Neurociencias de Rosario; Sanatorio Las Lomas; Hospital B. Rivadavia; Hospital Juan A. Fernández; Hospital Luis C. Lagomaggiore; Fundación Cerebro y Mente; INEBA; Centro “D. M. A. Moltedo”; Hospital de Agudos P. Piñero; Universidad Provincial del Sudoeste

**Author notes:** Corresponding author: Prof. Dra. Mariana Bendersky. There are no conflicts of interest.

**Keywords:** coronavirus, COVID-19, neurological, SARS-CoV2, nervous system

## Abstract

COVID-19 disease has spread around the world since December 2019. Neurological symptoms are part of its clinical spectrum.

**Objective:** To know the neurological manifestations in patients infected by COVID-19 in Argentina.

**Methods:** Multicenter study conducted in adults, from May 2020 to January 2021, with confirmed COVID-19 and neurological symptoms. Demographic variables, existence of systemic or neurological comorbidities, the form of onset of the infection, alteration in complementary studies and the degree of severity of neurological symptoms were recorded.

**Results:** 817 patients from all over the country were included, 52% male, mean age 38 years, most of them without comorbidities or previous neurological pathology. The first symptom of the infection was neurological in 56.2% of the cases, predominantly headache (69%), then anosmia / ageusia (66%). Myalgias (52%), allodynia / hyperalgesia (18%), and asthenia (6%) were also reported. 3.2% showed diffuse CNS involvement such as encephalopathy or seizures. 1.7% had cerebrovascular complications. Sleep disorders were observed in 3.2%. 6 patients were reported with Guillain Barré (GBS), peripheral neuropathy (3.4%), tongue paresthesia (0.6%), hearing loss (0.4%), plexopathy (0.3%). The severity of neurological symptoms was correlated with age and the existence of comorbidities.

**Conclusions:** Our results, similar to those of other countries, show two types of neurological symptoms associated with COVID-19: some potentially disabling or fatal such as GBS or encephalitis, and others less devastating, but more frequent such as headache or anosmia that demand increasingly long-term care.

## Introduction

At the end of 2019, a new coronavirus, SARS-CoV-2, spread rapidly throughout China and since January 2020 it was considered a pandemic by the WHO^1^. A little more than a year later, more than one hundred million cases and two million deaths have been reported worldwide: in Argentina, more than two million cases and almost 50,000 deaths^2^. It is the largest and most severe pandemic since the 1918 flu^3^.

Clinically, it is characterized by a mainly respiratory compromise of variable severity, mainly in subjects with advanced ages and pre-existing diseases. It ranges from a mild and self-limited disease to acute respiratory distress syndrome (SARS), a complication with a high mortality. As the pandemic progresses, reports of associated neurological manifestations are increasingly frequent. The rates vary according to the study methodology and the characteristics of the patients, but seem to occur in approximately 1/3 of the cases, which on the scale of the actual pandemic means a large number of people.

Virtually any part of the neuroaxis appears to be susceptible to be injured by SARS-CoV-2. As in other viral infections, the compromise could be due to direct effects of the virus, to the systemic response to the infection, to a combination of complications of systemic diseases, to para- or post-infectious inflammation, or later in the form of demyelinating lesions^4^. Hypoxemia, in patients with severe COVID-19, is likely to play a role in the development of encephalopathy, as do metabolic disorders due to organ failure and the effects of medication. A series of neuropathological cases of encephalopathic patients with COVID-19 revealed acute hypoxic ischemic damage in all^5^. In other series, neuroimaging findings were consistent with advanced post-hypoxic leukoencephalopathy, similar to those described in patients with other causes of SARS^6,7^.

Renin-angiotensin system dysfunction is another pathophysiological mechanism of COVID-19 infection. SARS-CoV-2 uses angiotensin-converting enzyme 2 (ACE2) to enter cells (Figure 1a). ACE2 converts angiotensin (AT) II to AT, which has vasodilator, anti-proliferative, and anti-fibrotic properties. By binding to ACE2, the virus causes dysfunction of this system, with cardiovascular and cerebrovascular consequences.

**Figure 1.**
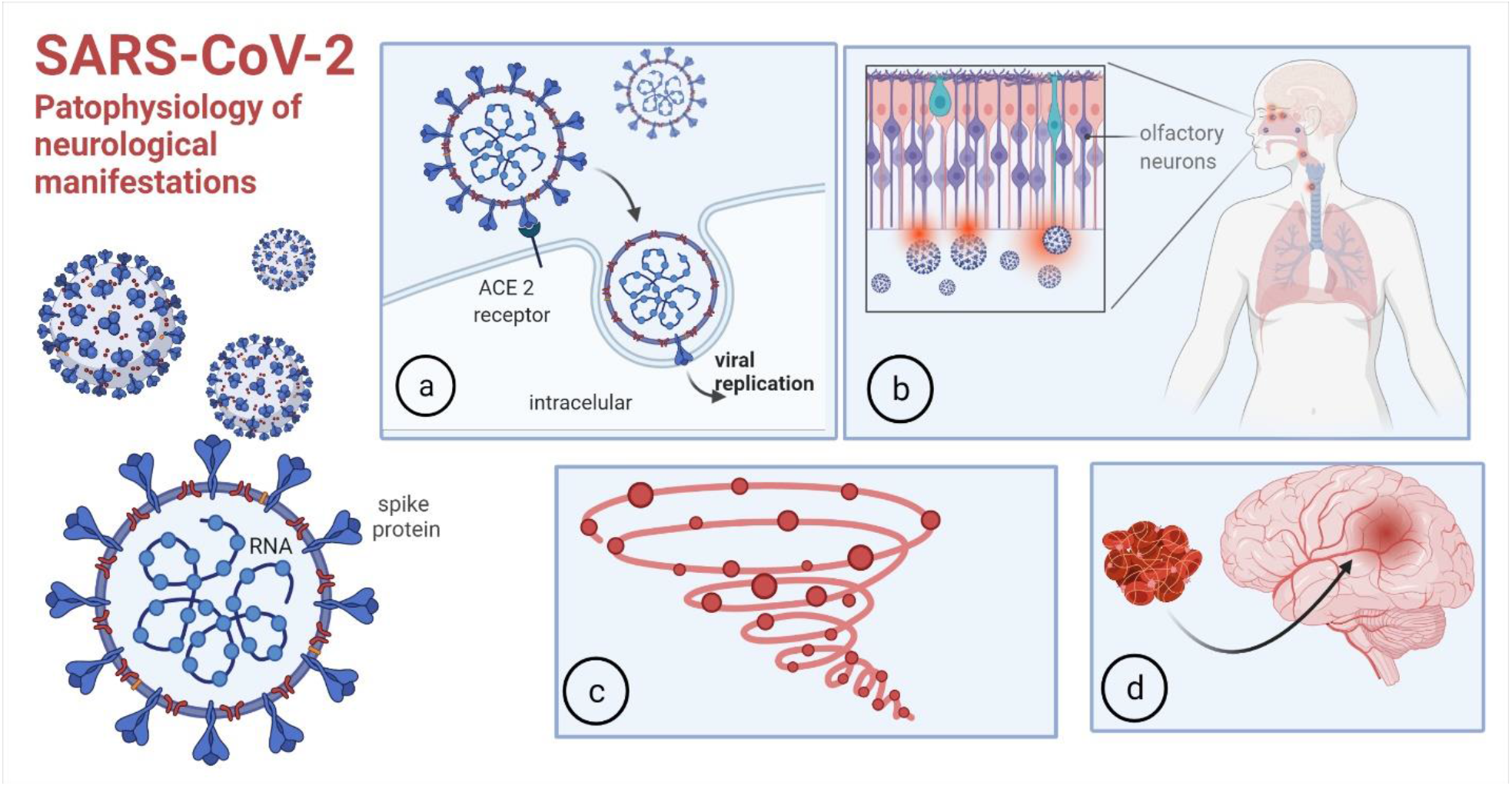
Pathophysiology of neurological manifestations. a-COVID-19 binding to ACE-2 to enter cells b-entry through the olfactory mucosa c-Dysregulated immune response, cytokine storm d-Thrombophilia-Created with BioRender.com.

SARS-CoV-2 can invade the brain through the olfactory epithelium and the neural-mucous interface (Figure 1b), and this causes anosmia in a high percentage of patients, often being the initial symptom. Later, it seems to spread by axonal transport and / or invasion of neighboring vessels, being able to reach the respiratory centers in the medulla oblongata, causing the irreversible respiratory failure of severe COVID-19, typically characterized by lack of dyspnea^8^. Some reports provide evidence of direct viral invasion of the nervous system: in a series of post-mortem cases, SARS-CoV-2 was detected in 53% of brain samples^9^.

There is also a dysregulated systemic immune response to the virus^10^. Critically ill patients with COVID-19 often develop signs of severe systemic inflammation similar to the cytokine storm syndrome (Figure 1c) with persistent fever and elevated inflammatory markers. This can cause altered consciousness *per se*, and can be associated with thrombophilia^11^, increasing the risk of cerebrovascular accident (CVA) and other thrombotic events (Figure 1d). Complement activation can also cause thrombotic microvascular injury in severe COVID-19 patients.

Finally, the virus can trigger para- and post-infectious phenomena, such as Guillain Barre syndrome (GBS)^12^ or CNS demyelination.

To date, there are no studies that have focused exclusively, consecutively, and prospectively on patients with neurological manifestations in our region. From the Argentine Neurological Society this Registry was launched, in order to know the neurological manifestations in patients infected by COVID-19 in Argentina, and to collect epidemiological and clinical data that could be correlated with its appearance.

## Subjects and methods

Multicenter, analytical, cross-sectional study carried out in adult patients in Argentina, from May 2020 to January 2021. All neurologists in the country were invited to participate, and they had to complete an online form. The protocol was evaluated by the research ethics committees of each participating unit. To ensure the anonymity of the patients and avoid duplicate records, each patient was assigned an alphanumeric code. Neurological signs and symptoms were recorded by trained neurologists.

Consecutive subjects older than 16 years were included, who consulted both in person and online, with some neurological symptoms, and criteria for a confirmed case of COVID-19, according to the definitions made by the Ministry of Health of Argentina (https://www.argentina.gob.ar/salud/coronavirus-COVID-19/definicion-de-caso). Patients with severe respiratory infections with another proven etiology were excluded.

The following variables were considered: *Primary:* confirmed COVID-19. Presence of neurological signs or symptoms and / or exacerbation of pre-existing neurological symptoms. *Secondary:* a-Demographic: age, sex, place of residence b-Systemic comorbidities c-Previous neurological disease d-If the neurological sign or symptom was the beginning of the COVID-19 infection e-Abnormal laboratory data f - Abnormal complementary neurological studies g-Degree of severity of neurological symptoms.

Continuous variables were expressed as means and standard deviation, or median with ranges of interquartile values (IQR). Categorical data were expressed as absolute and percentage values. The age distribution was divided into subgroups (Figure 2). The proportions of the categorical variables were compared using the χ2 test. *Pearson, Spearman* or *Mann-Whitney* tests were used to correlate variables, depending on the case. A value of p≤0.05 was considered as statistically significant.

**Figure 2.**
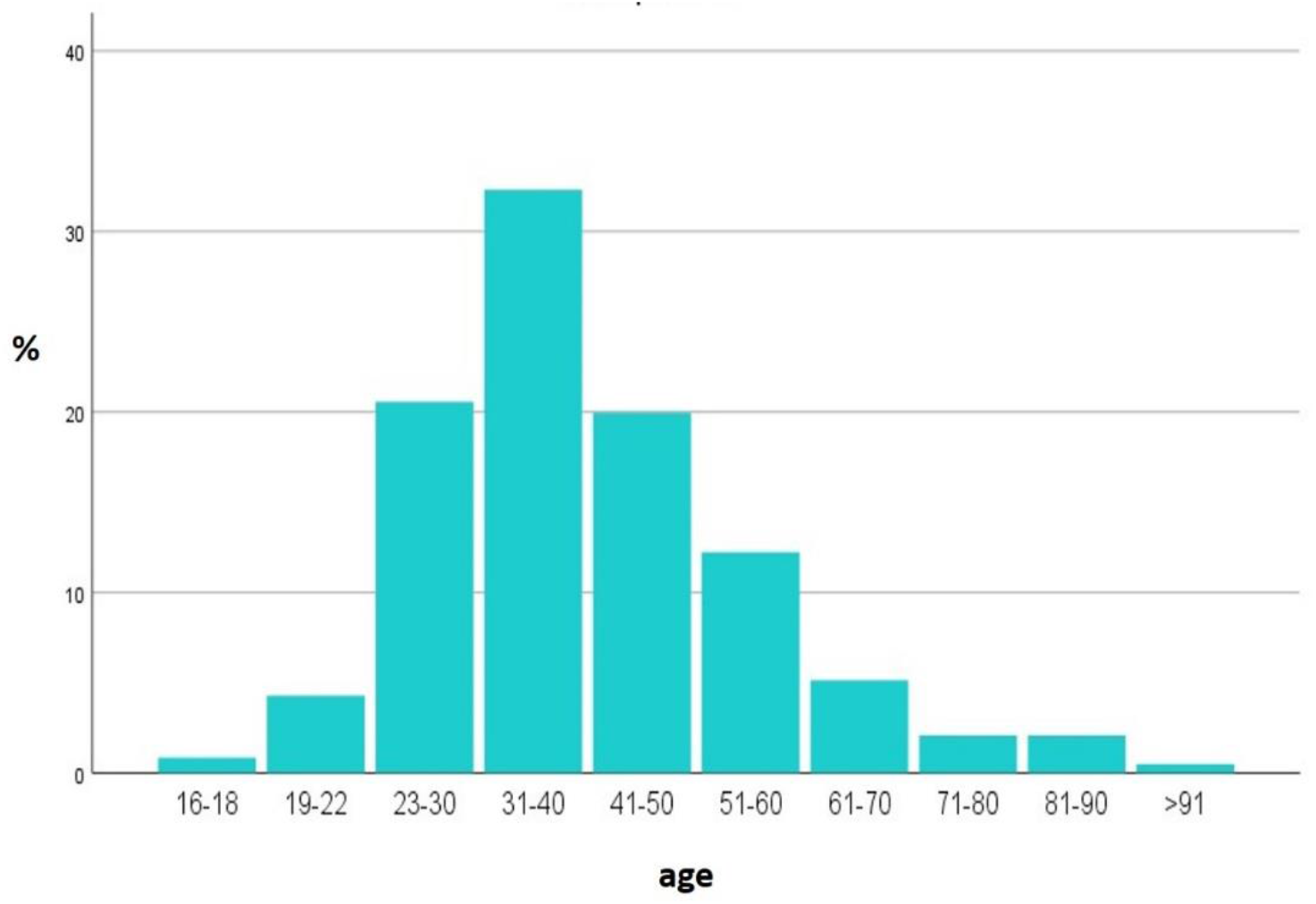
Age distribution.

## Results

The registry has 817 patients with neurological manifestations from different regions of the country, 52% men, 48% women, from 16 to 98 years old, average 38 (IQR 30-48) (Figure 2). The majority residents of Buenos Aires city (CABA) and surroundings (AMBA), in third place, Buenos Aires province, then the rest of the country, coinciding with the distribution of cases in Argentina (Figure 3).

**Figure 3.**
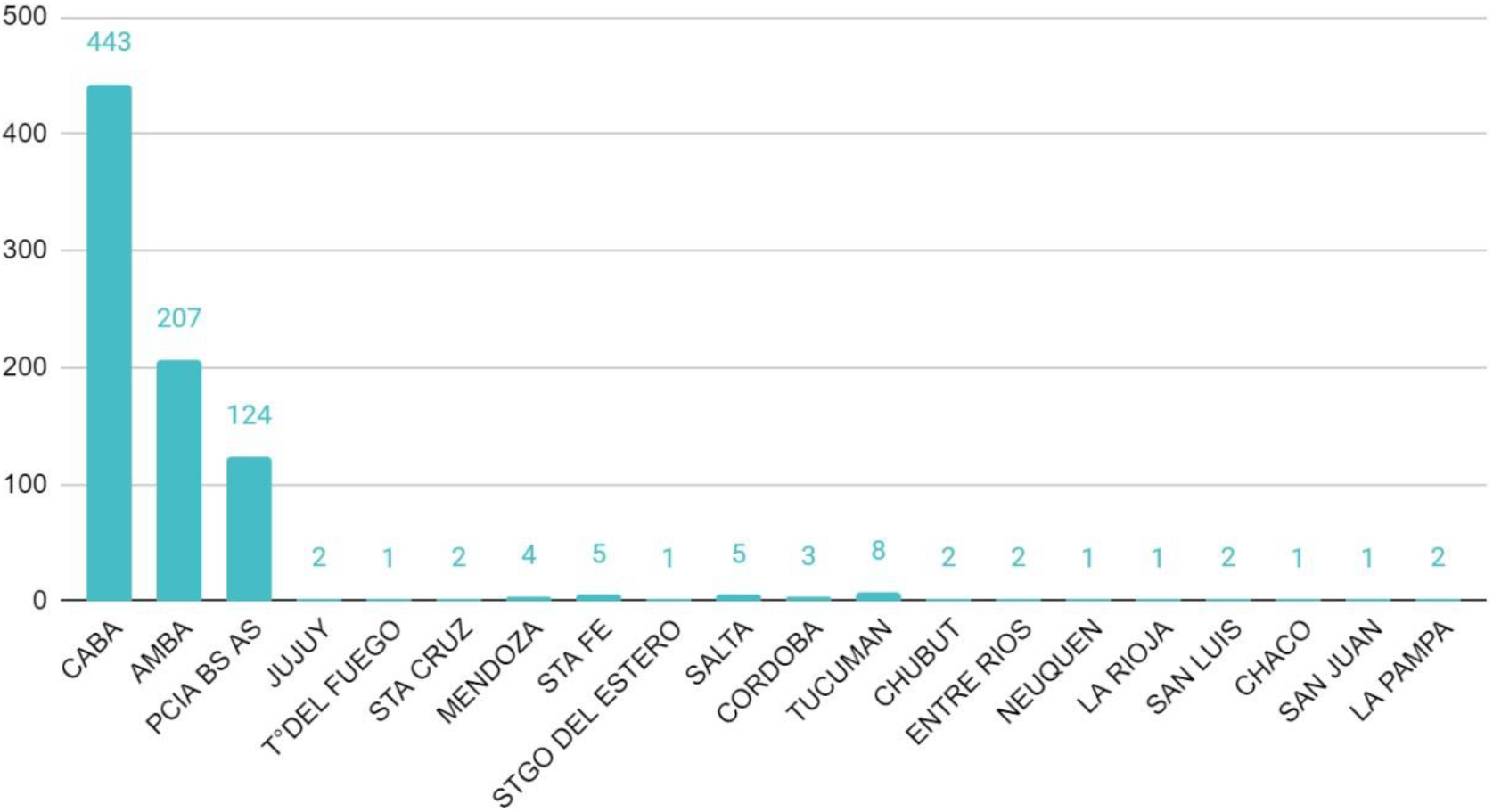
Place of residence of the patients.

69% of the subjects did not have any comorbidity, in the remaining, the most frequent was hypertension, followed by smoking or COPD (Table 1). Some reported more than one comorbidity (average 1.12).

**Table 1.** Comorbidities

|  | % |
| --- | --- |
| none | 68,9 |
| HTA | 12,0 |
| TBQ-COPD | 9,9 |
| Obese/overweight | 4,5 |
| cardiovascular | 4,0 |
| other | 3,7 |
| Diabetes | 3,6 |
| Dyslipemia | 2,4 |
| immunocompromised | 1,0 |
| VIH | 0,9 |
| pregnancy | 0,9 |
| oncological | 0,8 |

The majority (93.2%) had no previous neurological pathology (Table 2). This registry consists mainly of essential workers, due to the quarantine measures in force in our country during the considered period. Patients with moderate or severe neurological diseases were usually isolated and particularly attentive to COVID-19 prevention measures, so the prevalence of severe pre-existing neurological pathology was low in this registry.

**Table 2.** Previous neurological disease

|  | N | % |
| --- | --- | --- |
| None | 762 | 93,2 |
| migraine | 26 | 3,2 |
| vascular | 8 | 1,0 |
| demyelinating | 9 | 1,1 |
| movement disorder | 4 | 0,5 |
| dementia | 7 | 0,9 |

Regarding the neurological manifestations (Table 3), the most frequent was headache (69%), with photophobia (2.4%), retroocular pain (3.1%) or meningeal signs (0.9%). Second, anosmia / ageusia (66%). Myalgias (52%), and other painful manifestations such as allodynia or hyperalgesia (18%) were also reported. 6% referred asthenia. 3.2% showed signs of diffuse CNS involvement, such as encephalopathy (2%), agitation (1%), seizures (0.4%), pyramidal signs (0.2%), dysexecutive syndrome (0.5%), myoclonus (0.3%) or syncope (0.4%). 1.7% had cerebrovascular complications: ischemic stroke (1.3%) or hemorrhagic (0.4%). Sleep disorders were observed in 3.2%: insomnia (2.8%), or parasomnias (0.4%). Among the complications of the peripheral nervous system (in addition to anosmia), 6 patients with GBS (0.8%), 1 facial paralysis (0.1%), hearing loss (0.4%), plexopathy (0.3%), peripheral neuropathy (3.4%), tongue paresthesias (0.6%), were reported. On average, each one reported 2.35 symptoms. Compared with men, women reported a higher proportion of headache (p = 0.0036), vertigo (p = 0.0217) lingual paresthesias (p = 0.0211) and worsening of previous neurological diseases (p = 0.005). The rest of the neurological manifestations did not show differences by sex.

**Table 3.** Neurological signs and symptoms, grouped by categories (see detailed percentages in the text)

| **Signs and symptoms** | **N (%)** |
| --- | --- |
| Headache, meningeal syndrome (photophobia, retroocular pain, meningeal signs) | 556 (71%) |
| anosmia/ageusia | 519 (66%) |
| Pain syndromes (myalgias, allodynia, hyperesthesia, neuralgia) | 430 (55%) |
| PNS compromise (GBS, poli or mononeuropathy, myopathy, hearing loss) | 72 (9.2%) |
| Diffuse CNS involvement (Encephalopathy, seizures, agitation, myoclonus, dysexecutive syndrome, visual disturbances) | 25 (3.2%) |
| Sleep disorders | 25 (3.2%) |
| Worsening of previous pathology | 16 (2.0%) |
| stroke | 13 (1.7%) |

The first symptom of the infection was neurological in 56.2% of the cases. Most of them (66.24%) were of mild intensity, 30.42% of moderate intensity, and the remaining, severe. (Figure 4) Individuals with severe manifestations were older than those with mild or moderate manifestations (50.5 +/- 15.53 vs. 40.52 +/- 14.9 years; p = 0.0009). The severity of neurological symptoms correlated positively with the existence of comorbidities (Rho = 0.879, p = 0.004). Women presented a higher proportion of symptoms classified as serious than men (5.94 vs. 2.56%, p = 0.0214).

**Figure 4.**
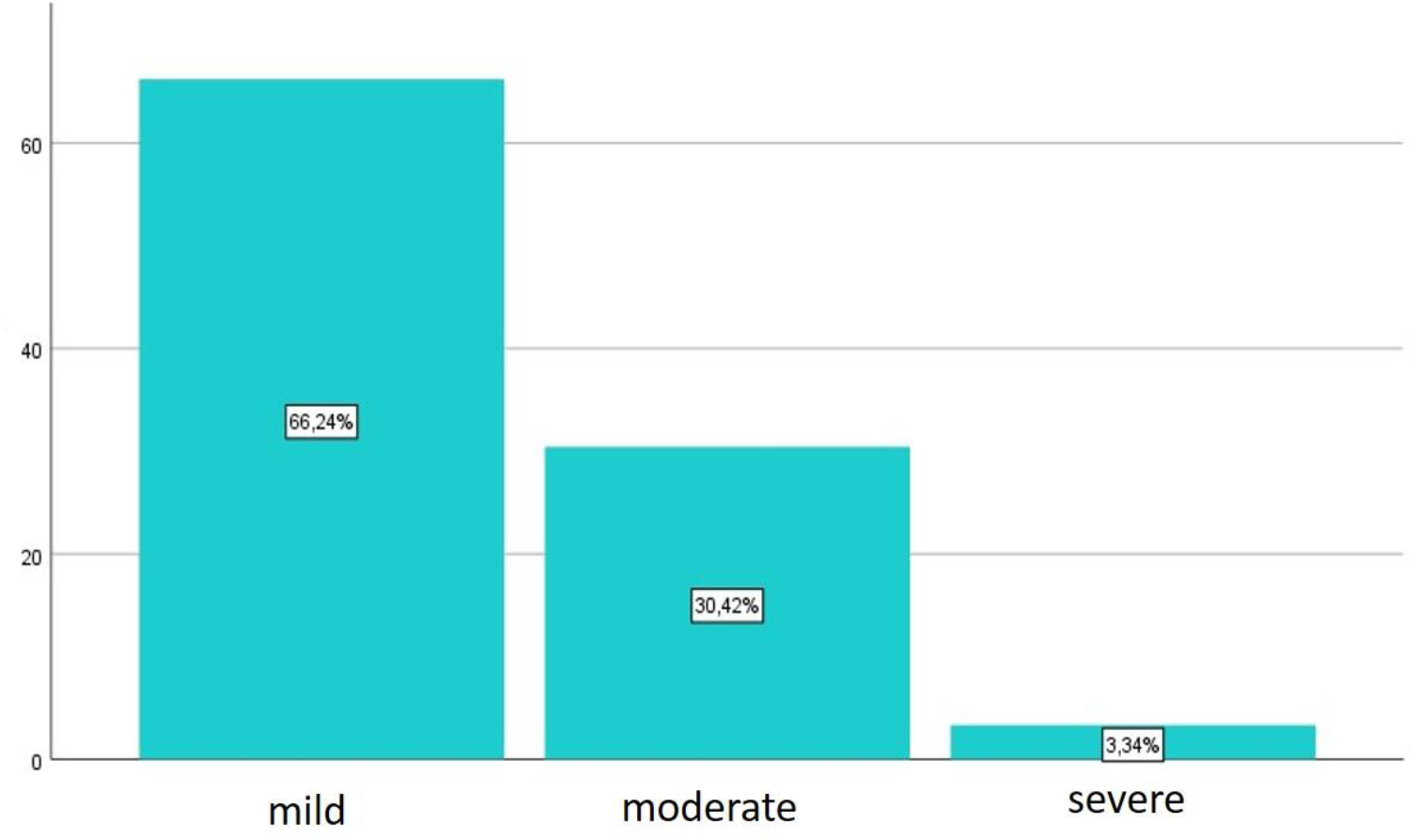
Severity of neurological manifestations.

Complementary studies were not carried out in all cases since the sample included patients from the community with home isolation, and only essential neurological studies were indicated in hospitalized patients. 31% of the patients had an abnormal finding: C-reactive protein (19%), D-dimer (10%), ferritin (9.4%), ESR (9.3%), LDH (9.5%), neuroimaging (1.7%), EMG (1.02%), EEG (0.77%) among others (Table 4).

**Table 4.** Abnormal complementary studies

| Characteristic | N = 777 (%) |
| --- | --- |
| Abnormal laboratory | 31 |
| PCR ↑ | 19 |
| Ddimer ↑ | 10 |
| LDH↑ | 9.5 |
| Ferritin ↑ | 9.4 |
| VSG ↑ | 9.3 |
| Transaminases ↑ | 5.9 |
| Lymphocytes ↓ | 5.9 |
| Abnormal renal function | 3.2 |
| Platelets ↓ | 2.3 |
| Eosinophils ↓ | 1.5 |
| CK ↑ | 1 |
| Albumin ↓ | 0.5 |
| CSF proteins ↑ | 0.4 |
| Fibrinogen ↑ | 0.4 |
| Anemia | 0.4 |
| Leukocytes ↑ | 0.3 |
| MRI-CT | 1.7 |
| EMG | 1.02 |
| EEG | 0.77 |

## Discussion

The first study that described the neurological manifestations of SARS-CoV-2 was carried out in Wuhan, and reported neurological involvement in 36.4% of the patients, with involvement of the CNS, PNS, and skeletal muscle^13^. Their results were similar to ours: headache, hyposmia and hypogeusia predominated. The greatest severity was correlated with advanced ages and pre-existing diseases, mostly hypertension. Patients with severe SARS-CoV2 had a higher incidence of stroke, encephalopathy, and muscle injury. In this subgroup, neutrophilia with lymphopenia and thrombocytopenia and other inflammatory markers, such as C-reactive protein and D-dimer, were observed. Later, more reports of neurological manifestations appeared in other countries^14–23^. For example, the Spanish registry included 841 hospitalized patients with COVID-19, with an average age well above that of our population, of which 57.4% developed some neurological symptom. Neurological complications were the main cause of death in 4.1% of all the deceased in the study^18^. An American study found that the independent risk factors for developing *any* neurological manifestation were severe COVID-19 and a younger age. 71.1% of the patients had a favorable functional outcome at discharge. However, encephalopathy was independently associated with poorer functional outcome and higher mortality within 30 days of hospitalization.^17^.

The late arrival of the pandemic in our region allowed us to record data from the beginning and to have local data, different from those of populations with other social and epidemiological contexts. For example, most of our patients had a mean age lower than that of the European population, and no comorbidity; even so, neurological manifestations were frequent. We found a higher frequency and severity of headaches and anosmia than those reported in other studies, often being the initial symptom; on the contrary, the prevalence of stroke associated with COVID was lower than that reported in other countries. We also found a frequent symptom, such as hyperalgesia / allodynia, rarely reported in other series of patients.

### Regarding some reported symptoms

#### Headache

It is a main symptom of acute SARS-CoV-2 infection and can become a persistent complaint of prolonged illness, even in mild cases. During the acute phase, the pain is throbbing or oppressive, worsens when bending over, and may be associated with photophobia and phonophobia. It can be located anywhere on the head, but it is mostly occipital and retroocular. Ding et al.^24^ describe headaches in 40% of the patients, of mild intensity, but in our series the percentage was higher, in many cases of moderate to severe intensity and with great functional repercussion.

Migraine-like characteristics may reflect an activation of the trigeminal-vascular system by inflammation or direct involvement of the virus. It has been reported that headache may be more common in patients with gastrointestinal symptoms, and that rapid weight loss with reduced appetite is characteristic of headache due to COVID-19^25^. In the chronic phase, pain can become constant. This chronic post-viral headache was recognized as a distinctive syndrome in 1986^26^. The 1890 pandemic, known as the “Russian or Asian flu,” which killed a million people, has extensive documentation on neurological sequelae that occurred months or years after the pandemic^27^, among them the “neurasthenia” syndrome: headache, lethargy and insomnia^28^, what we now know as a possible branch of chronic fatigue syndrome / myalgic encephalomyelitis and / or fibromyalgia, but some of these cases appear to be new persistent postinfectious daily headaches. The same symptom complex is now seen in patients with the long-term disease variant of SARS-CoV-2.

#### Anosmia/ ageusia

Sometimes an initial symptom, other times reported during the first week of infection, sudden anosmia without rhinorrhea is a striking and characteristic manifestation of SARS-CoV2. The abundance of reports of anosmia due to COVID-19 does not occur in other viral diseases. Many also have dysgeusia or ageusia (alteration or loss of taste, respectively), or changes in chemoesthesia (the ability to perceive irritants). The presence of viral RNA and proteins has been demonstrated in anatomically distinct regions of the nasopharynx and brain^9^. COVID-19 can enter the CNS by crossing the neural-mucosal interface in the olfactory mucosa, taking advantage of the close neighborhood of endothelial tissue and olfactory and trigeminal nerve endings. Abnormalities in the MRI signal were described in the olfactory bulbs of patients with COVID-19, which resolved in the follow-up^29,30^. In two autopsy cases, inflammatory infiltrate and axonal lesion were found in the olfactory tracts, but it was not possible to determine whether direct viral damage was responsible^16^. In a study carried out in hamsters, they suggest that COVID-19 anosmia would be related to a massive and rapid desquamation of the olfactory epithelium. They observed that two days after direct virus inoculation, half of the supportive cells were infected, but not the olfactory neurons^31^. The olfactory epithelium was completely shed, and the cilia had disappeared, with partial restoration 14 days later. In humans, the recovery of patients with anosmia is progressive, going from total anosmia, which lasts an average of one week, to hyposmia that resolves in approximately 2 weeks in most cases^32^.

The mechanism by which the virus causes loss of taste has not been fully explained. Flavor receptors detect chemicals in saliva and send the signal to the brain, but they do not contain ACE2, so they are unlikely to be infected with COVID-19. The sense of taste is transmitted by the glossopharyngeal, facial and vagus nerves, and only identifies basic flavors (sweet, salty, sour, bitter and umami); However, to recognize more complex flavors (the nuances of what is being consumed), the olfactory nerve is necessary. Although the taste seems to disappear with anosmia due to the latter, many people with SARS-CoV-2 develop a true and persistent ageusia. In addition, they report loss of perception of chemical substances. These sensations are not flavors, but their detection is transmitted by the trigeminal nerve, which does not express ACE2. The patients who recovered from the taste disorder first identified basic flavors, salty being the most reported, then sweet, sour and bitter^32^. Additionally, most of the patients manifested an independent recovery of smell, faster and more complete. All this could indicate the existence of an independent pathophysiology that affects different nerve pathways.

#### Myalgias

Myalgias, especially of the lower limbs and back, are common in viral infections. These discomforts occur immediately before or at the beginning of respiratory manifestations. Some speculate that it may be viral myositis, but conclusive evidence of this is lacking^33^. In Wuhan, 11% of patients had evidence of muscle injury with elevated creatine kinase (CK)^13^, in contrast, myalgia with normal CK was a common symptom in almost all patient series. Three case reports have described rhabdomyolysis with CK> 12,000 U / L. In one case, muscle biopsy in a patient with COVID-19 and myopathy showed perivascular inflammation and deposition of myxovirus-resistant protein A, a type I interferon-inducible protein ^34^.

#### Allodynia, hyperesthesia and other painful manifestations

An interesting symptom, probably underdiagnosed, reported in 16.9% of patients, is the presence of allodynia ^35^ (pain elicited by non-nociceptive stimuli) or cutaneous hyperesthesia (increased sensitivity to stimuli)^36^. An additional 0.8% reported paresthesias, without neuropathy. Physiopathologically, it is clear that SARS-CoV-2 infection is characterized by the appearance of a cytokine storm that leads to macrophage activation^37^. Cytokines are important in the development of allodynia and hyperalgesia in various pathological pain conditions, *hyperalgesic priming*. Another hypothesis suggests that neuroinvasion would affect ACE2-expressing neurons and microglia of the spinal cord. Since the binding of the spike protein to ACE2 causes a decrease in its levels in both cell types, the levels of angiotensin 2, which has pro-inflammatory and pro-algaeic effects, would increase. SARS-CoV-2 infection could also lead to a state of hyperstimulation resulting in autoimmunity, mitochondrial dysfunction and neuroinflammation, which could result in the development of fine fiber neuropathy. On the other hand, many patients recovering from SARS-CoV-2 infection show signs of critically ill polyneuropathy. Finally, prolonged administration of opioids in the intensive care unit may induce a state of increased pain sensitization, leading to allodynia and hyperalgesia along with myalgias and abdominal pain *(opioid-induced hyperalgesia*). Therefore, there are several reasons why COVID-19 patients are prone to developing neuropathic pain ^37^.

#### Neuromuscular complications

In our country, 6 cases of GBS syndrome were reported, which seems to be a rare complication. Among approximately 1,200 COVID-19 patients admitted for a month to three hospitals in Italy, only 5 GBS cases were identified^38^. The interval between the onset of viral disease and the development of muscle weakness is 5-10 days, similar to that seen for other GBS-associated viral infections. Some reports suggest that symptoms appear to progress more rapidly and be more severe than is typical for GBS; In one series, 3 out of 5 patients required mechanical ventilation, however, it was difficult to distinguish respiratory failure due to GBS from that due to COVID-19 lung disease. Dysautonomic characteristics were not observed in any of the series.

#### Stroke

Stroke appears to be relatively rare in the context of COVID-19. Rates of cerebrovascular events associated with COVID-19 are largely based on observational cohort studies in COVID-19 patients at different epicenters around the world ^10,13,15,18–23,39–43^, reflecting variable populations in terms of disease severity, comorbidities, and follow-up periods, this explains the variability in the rate of events. The risk of stroke can range from <1% in patients with mild SARS-COV2 to 6% in patients in intensive care. The incidence in this registry was 0.6% hemorrhagic stroke and 1% ischemic stroke.

In general, stroke occurs one to three weeks after the onset of COVID-19 symptoms. It can be ischemic, hemorrhagic, or present as venous thrombosis. Ischemia can also manifest as non-focal deficits, such as encephalopathy, and affect multiple vascular territories. They are usually cryptogenic, but can also be attributed to great vessel thrombosis, which can be simultaneous in different arterial territories, cardiogenic embolism or arterial dissection^44^. In patients under 50 years of age, a more severe initial neurological picture has been observed, in the context of severe SARS. The mortality of stroke in COVID patients is higher than usual and this is generally determined by the underlying disease. SARS-CoV-2 has been suggested to be associated with a hypercoagulable state. This is reflected in the elevated levels of D-dimer (marker of thrombus turnover) observed in many patients during the first weeks of the disease, particularly in the most severely affected^39,44^. These elevated levels of D-dimer are also present in some patients with ischemic stroke. Unusual cases of aggressive aortic, carotid and basilar thromboses have been reported^42,43,45^. There may also be a higher rate of early arterial reocclusion after mechanical thrombectomy^46^. SARS-CoV-2 has also been associated with several cardiac manifestations, such as arrhythmia, heart failure, and myocardial infarction, many of which can predispose to cardioembolic stroke. This heterogeneity suggests that the mechanisms of stroke in COVID-19 are multiple and may include both virus-specific pathophysiological characteristics and nonspecific effects of inflammation and coagulation dysfunction, superimposed on pre-existing risk factors.

#### Encephalopathy / agitation / seizures

Encephalopathy is common in critically ill patients with COVID-19. In a series of 58 patients with SARS related to COVID-19, encephalopathy was present in 2/3 of the patients^23^. In a study of 509 hospitalized patients with COVID-19, 31.8% had encephalopathy, and those patients were older than those without it, had a shorter time from symptom onset to hospitalization, were more likely males and were more likely to have comorbidities^17^. Sensory impairment can be the main symptom of COVID-19. In a study of 817 older patients (mean age 78 years) evaluated in the emergency department who were diagnosed with COVID-19 infection, encephalopathy was present in 28%. Among those patients, 37% lacked typical COVID-19 symptoms, such as fever or dyspnea ^47^.

The etiology of encephalopathy in these patients is usually multifactorial. Critically ill patients with COVID-19 are subject to the same causes of toxic-metabolic encephalopathy as other critically ill patients. Pyramidal signs are frequent, seizures may occur, they generally have no evidence of brain swelling on MRI or CSF analysis. The EEG shows nonspecific findings (diffuse bifrontal slowing). Bilateral frontal temporal hypoperfusion has been reported in some encephalopathic patients with SARS-CoV-2, 33% of the survivors showed a dysexecutive syndrome, with alterations in attention, orientation and ideomotor praxis, suggestive of frontal lobe involvement^23^.

In our registry, 1.5% of the patients were encephalopathic, 1% had agitation, and 0.5% had seizures. Only one patient had a dysexecutive syndrome. This low prevalence may be due to the significantly younger ages of our patients, the majority without comorbidities, and the probable underreporting of UCI patients.

Several limitations inherent in the results of this work must be considered. In the first place, there was under-registration of cases, since many patients did not arrive at the neurological consultation, being attended by general internists or ICU doctors. In addition, mild neurological symptoms may go unnoticed in patients with very severe disorders. Second, we cannot know what percentage of all patients infected by COVID-19 developed any of these manifestations. Based on data from the National Health Surveillance System ^48^, the National Ministry of Health reported that neurological symptoms occurred in 47.2% of all cases. In general, the proportion of patients with neurological manifestations is small compared to that of respiratory disease. However, the pandemic continues, with new and more contagious mutations of the virus. Projections that between 50% and 80% of the world’s population may be infected before herd immunity develops suggest that the total number of patients with neurological diseases could increase.

## Conclussions

This work seeks to raise awareness in the neurological community about the different neurological manifestations of COVID 19, in order to closely monitor them and avoid late or erroneous diagnoses. Our results, similar to those of other countries, show that COVID-19 exhibits two types of neurological symptoms: some potentially disabling or fatal such as GBS or encephalitis, and others less devastating such as headache, anosmia, and myalgia. These so-called *minor symptoms* in some cases continue for ≥12 weeks and have been labeled as *long COVID* or *post-COVID syndrome*^49^. Longitudinal neurological evaluations after recovery will be crucial in understanding the natural history of SARS-CoV-2 in the nervous system and monitoring possible sequelae.

Neurological involvement by COVID-19 is an issue of great importance that requires a global effort to obtain the epidemiological and clinical data necessary to quantify the magnitude of the problem, define the full range of neurological disease and promote neuropathological, pathophysiological and therapeutic research^41^. Neurological complications, both the most severe, which cause a lifelong disability, and the milder but much more frequent ones, increasingly demand long-term care, with health, social and economic costs that should be considered within future health plans and policies.

## Data Availability

all data referred in the manuscript are available upon request

